# Increasing Awareness of the Importance of Reporting Adverse Drug Reactions of Antiretroviral Drugs Among Adults Living with HIV in Moshi, Tanzania: A Pilot Study on Using SMS Reminders for Reporting

**DOI:** 10.1101/2023.04.04.23288072

**Authors:** Lyidia Masika, Naomi Emmanuel, Tumaini Mirai, Gloria Nyanungu, Mary Shirima, Marion Sumari–de Boer, Rehema Maro, Benson Mtesha, Kennedy Ngowi

## Abstract

**Introduction:** In Sub-Saharan Africa, there are knowledge gaps on adverse drug reactions (ADRs) of antiretroviral treatment (ART). Studies have shown that limited training capacity among healthcare workers has affected reporting of ADRs and weakened the pharmacovigilance system in Tanzania. Studies have shown that the use of digital tools, including SMS communication, might be a viable way to increase knowledge and understanding of reporting ADRs. This study aimed to examine the acceptability and feasibility of SMS texts to increase awareness about the importance of reporting ADRs of ART among people living with HIV (PLHIV) in Tanzania. We also explored different ADR profiles that were reported by the participants.

**Methodology:** This was a prospective implementation pilot study. PLHIV who consented to the study received a biweekly message to ask them how they were doing. We programmed keywords in the system that could be used for responding to the presence of ADR. The response to messages generated a flow of SMS that determined the presence of ADRs and informed participants to report. The technical feasibility was calculated based on the percentages of SMS which were sent and delivered about ADR and acceptability was measured based on the percentage of SMS that was replied to by the participants. We also collected data on different types of ADR reported by the participants. We conducted focus group discussions with participants and in-depth interviews with health workers to understand their experiences with and acceptability of the system.

**Results:** Over a 12-month study period, a total of 92 participants were recruited. Sixty-two (67.4%) were women. The mean age of our participants was 42 years (SD± 12). Forty (43.5%) participants were on ART for less than 3 months and the other 52 (56.5%) participants were on first-line ART. The system successfully sent and delivered 105 follow-up messages to all participants who consented to receive SMS. Among all the sent SMS 100 (95.2%) were replied to by the participants. Commonly reported ADRs were “itching”, gastrointestinal discomfort”, “joint pain”, “malaise” and “headache”. The themes identified through FGD and IDI were: high motivation to report ADR, well-understood SMS content, the system to remain the same, no concerns about unwanted disclosure, and difficulties in replying to SMS due to network. The latter was mentioned by a few participants.

**Conclusion:** SMS for creating awareness on ADRs reporting is a user-friendly intervention and highly accepted based on qualitative data among PLHIV in Tanzania. Also use of SMS is a low-cost intervention and a simple way to improve public health issues with society.

## Introduction

Human immunodeficiency virus (HIV) still causes high morbidity and mortality. In Tanzania, 52% of people living with HIV (PLHIV) aged 15-65 know their status and 91% of them are on antiretroviral treatment (ART)(1). ART must be taken every day since it does not provide a cure for the disease as it only suppresses viral growth(2). Despite the good outcomes, ART causes Adverse Drug Reactions (ADR), which are a burden to both the health system and the individual and may compromise the effectiveness of the ART programs(3). ADR is defined as any response to medication that is noxious and unintended that may occur at normal doses(4).

ADRs may range from mild to acute or even life-threatening. These reactions are individualized and they may not appear to everyone on certain medications due to different factors such as genetic differences (3,5). In addition, many people living with HIV are TB co-infected or have other co-morbidities such as diabetes and cardiovascular conditions; hence, they are exposed to both ART and anti-TB drugs or other drugs which may cause the risk of hepatotoxicity and renal damage to be higher (6,7). Data on ADR could inform policy guidelines on the best regimens for ART and this data should come from national pharmacovigilance (PV) systems (8–10).

Currently, the passive way of collecting data through national PV systems is not adequate (10). In Tanzania, all reactions, whether predictable or non-predictable, serious or mild, and known and unknown are reported as suspected ADRs and Adverse Events using special forms. However, according to Tanzania Medicine and Medical Devices Authority (TMDA) reports and other literature, only one report per one million inhabitants has been documented per year (9,10). This is contrary to the WHO average of reporting which is 200 reports per million people (11). Studies in South Africa, Tanzania, and Ethiopia have mentioned several challenges hindering spontaneous reporting of ADR including a lack of enforcement on sensitization of the importance of reporting ADRs in both health care staff and PLHIV (6,10,12).

Due to the high penetration of mobile phone use, the most common ways of communication in Tanzania are through phone calls and short message service (SMS) texts (13,14). Previous studies have shown that the use of digital tools, including SMS communication, might be a viable way to increase knowledge and understanding of reporting ADRs (15). In addition, it can remind all people involved in the disease management of PLHIV to report ADRs. Therefore, we designed an SMS system that triggered active reporting of ADRs in Tanzania. The overall objective of this study was to investigate the feasibility and acceptability of using SMS to improve awareness of the importance of ADR reporting. Specifically, for feasibility, we compared the trend of the number of SMS sent/delivered/failed and replied to among the study participants. Considering acceptability, we analyzed qualitative data on questions about the use of SMS to report ADR among the study participants. Furthermore, we also want to explore the different ADR profiles that are reported.

## Methods

### Study design

This was a prospective cohort pilot study conducted for 12 months using mixed-method approaches. The study was approved by the Kilimanjaro Christian Medical College Research Ethics and Review Committee (CRERC) and the National Health Research Ethics Sub-Committee (NatHREC) of Tanzania.

### Study area

Participants were enrolled from two HIV care and treatment clinics, which are Kilimanjaro Christian Medical Centre (KCMC) and Majengo Health center in Kilimanjaro region, Tanzania. KCMC is a zonal hospital for the Northern zone in Tanzania which has a care and treatment clinic for HIV serving more than 2000 clients per year. Majengo Health Centre is based in Moshi town and the clinic serves around 1000 clients.

### Study population

The study population included two groups: (1) people living with HIV and (2) healthcare workers who are serving PLHIV. The eligibility criteria for enrollment in the study of PLHIV were: age between 18 and 65, confirmed and documented HIV infection, patients on ARV treatment for less than 3 months more than 12 months, attending a CTC at KCMC or Majengo Health Centre, and able to read, understand SMS and write SMS. Individuals who were critically ill or participating in other trials were excluded from the study. Eligibility criteria for the Health Care Workers were: age between 18 and 65 years old and working at the CTC of KCMC or Majengo health center.

### Sample size calculation

As this was a pilot study, we used a convenient sample size of 100 PLHIV. Five healthcare workers were interviewed at the end of the study to collect data on their perceptions of the intervention.

### Study procedures

General information on the study was provided to all participants while they waited for consultation at the clinic. Those who were interested were invited to meet with the study nurse for more information. After detailed information was given by the study nurse, participants who agreed to participate in the study signed informed consent and were enrolled in the study if they met all inclusion criteria. During enrolment, patients received an extensive explanation about adverse drug reactions and what they are. This was to ensure that they knew what they will have to report.

#### SMS scheme

An SMS system was developed to send an automated bi-weekly SMS to participants. The messages were sent on Wednesdays every two weeks during the total study period. Once enrolled, participants received a welcoming message describing the importance of reporting any ADR. Then they received another follow-up SMS with the question asking “How are you doing and if they had any symptoms?”. Participants were required to respond to the questions with special characters that could be recognized by the SMS system. The characters included: 1. “I’m okay.” and 2. “I’m not okay”. If participants responded “2”, the system responded automatically with an SMS text stating the importance of reporting the symptoms and asking if they wanted to report the symptoms. If he/she replied with yes, the system asked to choose how they wished to report the ADR choosing from the following options: (1) Complete a web-based ADR reporting form, (2) Complete a green form at the clinic, (3) Receive a call from a health care worker for completing the yellow form together or (4) Visit the clinic/wait till next visit to complete the yellow form. A week later, a follow-up message was sent to ask if the symptoms were reported, and if answered with “yes”, which means of reporting was used.

### Data collection

#### Case report form

Instantly after enrollment, baseline data were collected using a case report form, which included demographic data including gender, age, education level, and phone number for receiving SMS. In addition, data on HIV-related characteristics such as type of ART taken, time of medication, and if there were any other medications taken were collected. Follow-up after enrollment was done for up to one year during the standard clinic visits. Participants were interviewed every time they visited the clinic on their routine HIV care and treatment. Information about those who reported through SMS to have an ADR was shared with the nurses for further follow-up.

#### Qualitative Data

After one year of follow-up, we conducted in-depth interviews with HCWs about their experience with the intervention. The interviews were done in the local language (Kiswahili) by a research assistant (RS). The topic guides were developed by the study team and included issues that surfaced during the study. The following topics have been explored: experience with the system, usefulness of the system, technical issues, ease and difficulties, disadvantages/Advantages, potential stigma, and appropriateness of the SMS. Additionally, focus-group discussions were organized with participants using the same topic list. Each FGD accommodated 8-10 participants with the following group characteristics: (1) women aged over 30, (2) women aged up to 30, (3) men aged over 30, and (4) men aged up to 30. The interviews and focus group discussions were recorded, transcribed, and translated by the same research assistant (RA).

#### SMS Data

We extracted the number of SMS sent and received including the follow-up SMS “Did you get any symptom after taking the drugs?”, SMS on reporting ADR “ Willingness of reporting the symptoms”, SMS response about ADR reporting “Preferred and means of reporting ADR”.

### Data analyses

Data analysis of quantitative data was conducted using STATA Version 15.5. Descriptive analysis was done to summarize the demographic characteristics of the study participants in frequencies and percentages. To investigate the technical feasibility of the SMS system, we calculated the frequency and percentage of total SMS successfully sent and delivered and the number of failed SMS. Furthermore, we analyzed the preference for ADR reporting by determining the number of follow-up SMS on ADR and responded SMS on willingness to report ADR as well as means of reporting.

To investigate the acceptability of the system among healthcare workers and participants, firstly KN, NE, and RM read and re-read all the transcripts and produced initials codes. Discussions to harmonize the initial codes were done among KN, NE, and RM to produce a final codebook. Transcripts were uploaded to MAXDQA for coding. Later, thematic framework analysis with an inductive content approach was applied to analyze and extract the quotes from the transcripts. To explore the difference in reporting ADRs between groups, we summarized the most commonly reported ADRs.

## Results

### Baseline characteristics of the study population

Over 10 months, 114 participants were screened for eligibility, of which 10 (9%) did not meet the eligibility criteria and 12(10%) declined participation. Among 92 participants enrolled in the study, 45 were from Majengo Health Centre and 47 were from Kilimanjaro Christian Medical Centre. The mean age of our participants was 42 years (SD± 12). Forty (43.5%) participants were on ART for less than 3 months and the other 52 (56.5%) participants were on first-line ART. Participants from KCMC hospital reported more ADR than those at Majengo HC, while participants on second-line treatment encountered more ADR than those in the first line (Table 1).

**Table 1.** Baseline characteristics of study participants and proportion of ADR reporting (N=92)

| Variable | N (%) | ADR REPORTING |
| --- | --- | --- |
| **Study Site** |  |  |
| Kilimanjaro Christian Medical Centre | 47 (51.1) | 35 (74.5%) |
| Majengo Health Centre | 45 (48.9) | 7 (15.6%) |
| **Age (Years)** |  |  |
| Mean(±SD) | 42 (12) |  |
| **Sex** |  |  |
| Female | 62 (67.4) | 29 (46.8%) |
| Male | 30 (32.4) | 13 (43.3%) |
| **Duration of ART treatment (months)** |  |  |
| Less than 3months | 40 (43.5) | 13 (30.9%) |
| More than 12 months | 52 (56.5) | 29 (69.1%) |
| **Education** |  |  |
| Primary | 53 (57.6) | 20 (47.6%) |
| Secondary | 30 ( 32.6) | 15 (35.7%) |
| Tertiary | 9 (9.8) | 7 (16.7%) |
| **Marital Status** |  |  |
| Single | 29 (31.5) | 16 (55.2%) |
| Married | 33 (35.9) | 15 (45.5%) |
| Separated | 20 ( 21.7) | 6 (30.0%) |
| Widowed | 10 ( 10.9) | 5 (50.0%) |
| **Antiretroviral Medications\*** |  |  |
| First line drugs | 77 (83.7) | 31 (40.3%) |
| Second line drugs | 14 (15.2) | 10 (71.4%) |

\*missing values frequency do not tally.

### Reported ADR profiles during the study period

Several ADRs were reported from different patients. The majority of the participants reported gastrointestinal discomfort including diarrhea. A few participants had complaints of Hiccups, elevated glucose levels, lack of sexual drive, and sore tongue after taking their medication (Table 2).

**Table 2:** Reported ADR profiles during the study period.

| Reported Cases | Frequency (N) |
| --- | --- |
| Gastrointestinal discomfort which included diarrhea | 12 |
| Skin rash | 11 |
| Difficulties in breathing | 5 |
| Chest tightness after taking the medications | 5 |
| Hallucinations and severe bad dreams | 5 |
| Dizziness | 5 |
| Headache | 5 |
| Numbness or jaundice or muscle pain or Sweats and general body malaise | 4 |
| Insomnia | 2 |
| Uneven fat distribution | 2 |
| Metallic taste and persistent thirst | 2 |
| Elevated blood glucose levels | 2 |
| Sore tongue | 1 |
| Lack of sexual drive | 1 |
| Hiccups | 1 |

#### Preference of route of ADR reporting

A total of 105 follow-up SMS were sent to participants in question if they encountered any ADR/a symptom from their medications. Seventy-one (67.6%) SMS replied that they had a symptom after taking medications. Among them, 62 (87.3%) were willing to report the symptoms, 3 (4.2%) did not wish to report the symptoms and 6 (8.5%) did not reply to the message on willingness to report ADR. A total of 42 (45.7%) participants reported will visit the clinic for reporting, 10 (16.1%) were called by the study nurse to report and 8 (12.9%) reported ADR through the website. (See table 3)

**Table 3:** Preference for ADR reporting.

| QUESTIONS | N | % |
| --- | --- | --- |
| **Did you get any symptoms after taking the drugs?** |  |  |
| YES | 71 | 67.6 |
| NO | 29 | 27.6 |
| NO REPLY | 5 | 4.8 |
| **The willingness of reporting the symptoms** |  |  |
| YES | 62 | 87.3 |
| NO | 3 | 4.2 |
| NO REPLY | 6 | 8.5 |
| **Preferred means of reporting** |  |  |
| Website | 37 | 59.7 |
| Green card | 9 | 14.5 |
| Call from Health Care Worker | 1 | 1.6 |
| Visit clinic | 4 | 6.5 |
| NO REPLY | 11 | 17.7 |
| **Means used to report** |  |  |
| Website | 8 | 12.9 |
| Green card | 0 | 0 |
| Call from Health Care Worker | 10 | 16.1 |
| Visit clinic | 42 | 67.7 |
| NO REPLY | 2 | 3.2 |

**Table 4:**
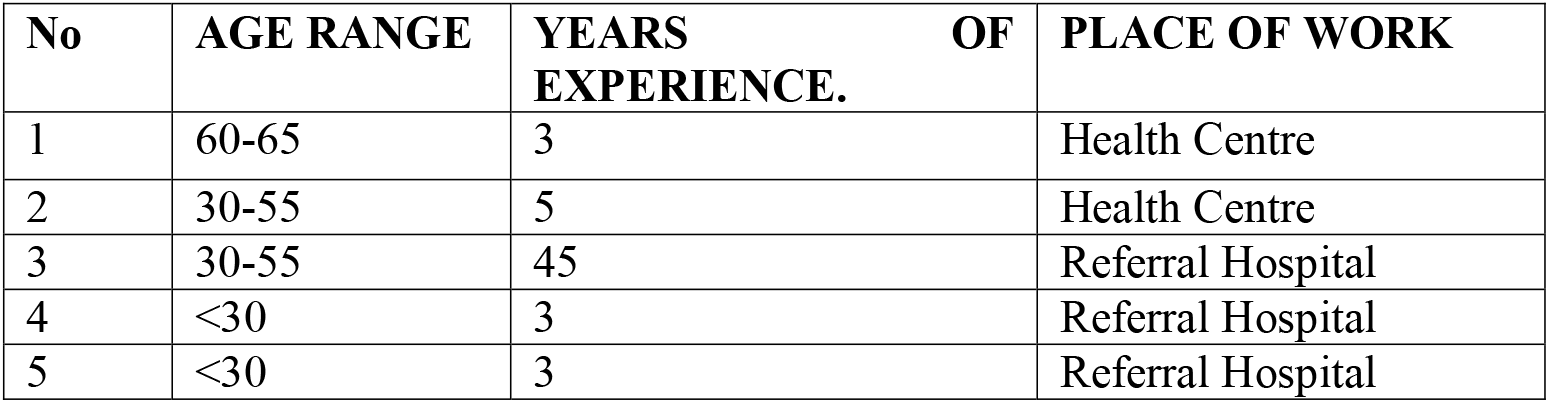
IDI- Healthcare providers.

| No | AGE RANGE | YEARS OF EXPERIENCE. | PLACE OF WORK |
| --- | --- | --- | --- |
| 1 | 60-65 | 3 | Health Centre |
| 2 | 30-55 | 5 | Health Centre |
| 3 | 30-55 | 45 | Referral Hospital |
| 4 | <30 | 3 | Referral Hospital |
| 5 | <30 | 3 | Referral Hospital |

**Table 5:** FDG-Participants.

| Variable | Number |
| --- | --- |
| Sex |  |
| Male | 7(26%) |
| Female | 20(74%) |
| Occupation | Entrepreneurs<br>Housewife<br>Cleaner<br>Farmer<br>Employee |
| Education | Primary<br>Secondary<br>University<br>No education |
| Age median | 46(17-64) |

### Qualitative results

Several themes came up during the IDI and FGD. Below, they have been described including quotes from participants to illustrate each theme.

#### General Experience and acceptability of the System

The general experience with the SMS system, including the SMS content and SMS frequency, was reported to be beneficial for reporting adverse reactions. For instance, on the topic of SMS content, the majority indicated that the contents were well understood, easy to follow, and effective to trigger them to report adverse reactions. Others described the content to be informative, particularly on means of reporting medication reactions. For instance, a **woman in her 40’s** said:

> “*Yes the SMS was very good. First, they reminded me to report if I have medication reactions, and second which means I should use it to report those events”*.

Another **woman added:**

> *“I did not have any problem with the SMS contents because the message didn’t state the word HIV or virus so it was okay. Also, if the system will continue in the future, don’t change anything in the REMIND-ADR system*.*”*

The frequency of the messages was another important positive experience described by the majority of participants. Almost all participants described being happy with the number of messages being sent by the system.

This was **described by a male in his 50’s:**

> “*I like the way the SMS was being sent with good information about how to report medication effects*.*”*

Also, he added:

> *“There was a time when the messages stopped being sent while I wished them to continue, I felt very bad*.*”*

Furthermore, many were comfortable with the system because the nurses had explained to them the importance of the system in reporting adverse reactions.

#### The usefulness of the system

Participants were first asked how useful the system was in the context of triggering them to report.

Most of them reported that the system was useful because they were motivated by the SMS and felt being cared about their well-being. Others indicated that the system should continue even after the end of the project. One participant described:

> *“I had my phone with me all the time so when the messages came I felt like being cared for by other people*.”

Correspondingly, in the IDI, the nurses declared that participants were satisfied with the system and it helped them to report adverse drug reactions. Also, the study increased awareness about drug reactions as well as communication between patients and nurses during clinic visits.

This was shown when the health care provider said:

> *“They were satisfied because so many patients were able to get the message, reply to it and help even those who experienced side effects of the drugs to be able to detect early*.*”*

Furthermore, reporting ADR helped nurses to monitor and counsel patients more closely. Also, the system was useful to provide extra information, which led to identifying new cases with ADR.

This was also supported by a healthcare provider who said:

> *“It helped me to realize that it is my responsibility to talk to the patient and advise them about medication and also to tell them that if they see any problems after starting medication they should report it*.***”***

Another **healthcare provider** revealed:

> *“I came across a new case like the ones I told you about, the one who says he doesn’t see, the other person doesn’t feel like having sex, and the man who got those hiccups. There was another we found. There was a woman was who had like a burn in her mouth. All these, we have never encountered before”*

#### Easy to Report

Participants were asked to describe the way the system works based on their own experience. The majority described that the flow of the SMS through the system was easy and well understood by participants. Most of the clients that got reactions indicated there were no difficulties in reporting as the instructions were easy to understand. As mentioned by **a male in his 50’s:**

> *“When I answer the question that I’m sick and I want to report the events, I was called by the nurse to go to the clinic*.*”*

**A female in her 40’s** said:

> “*Yes, I had to stop the medication and use fruits because the sleep was becoming too much and I was losing my strength, I reported this*.*”*

#### Potential barriers to using the SMS

Although the majority described the system as good, few themes emerged during the FGD discussions describing potential barriers. One of these barriers was the poor network in some areas, which led to a delay or even failure in replying. Another barrier was not having an SMS bundle to reply to the messages. Despite that SMS bundles were being provided by the study throughout, some described they did not get them. Hence, they were not able to reply.

A **male** in his 40’s said:

> *‘‘The number that I had registered as an [name of a network service provider] number. When I went to the village, I found no network. So, I was forced to have a sim card from a new provider*.*”*

Potential stigma was among the barriers discussed. Even though the majority owns a phone, one participant had concerns about not knowing when to expect the message because of sharing the phone with family members. A woman **in her 20’s** described:

> *“Sometimes I shared my phone with other people, so when the message came, I felt like exposing REMINDER SMS might expose my status. Therefore, sometimes I deleted messages without reading them*.*”*

Changing the phone number of some patients without informing their healthcare providers resulted in not receiving reminder messages for reporting adverse drug reactions on time. As one healthcare provider said:

> *“The patient was changing the number, so when you look for them on that number, you do not find them. So, you find the day you look for them again on the number maybe he receives. He says: ‘I changed the number*.*’ We ask him for the number he uses currently. We ask them not to change numbers, but now everyone has the things they can still change*.*”*

Some participants perceive responding to the SMS as a waste of time and a nuisance because they perceived it to have no benefits of it, though the nurses tried to advise them. This was explained by a healthcare provider who said:

> *“Sometimes I see answering the message every time won’t help me, I don’t even see any benefit despite the nurses advising that SMS were beneficial for my health and for other people as well*.*”*

Though the system was widely accepted some of the users wished the word “medication” to be changed to avoid unwanted disclosure. Also, others mentioned that the word “medication” is easy for someone who saw the message to know what kind of medication they are using. A **male in his 40’s said:**

> “*You know, the problem is the word ‘medication’ maybe if it is kept in the plural. When you put it in the plural, you already remove someone from the meaning. Like, do you have any problem with the medications that you are using, simple and clear? Someone won’t understand the meaning. Since it might be aspirin or chloroquine. So it’s just there in the singular and plural. They should write medication instead of medications*.*”*

Few reported having challenges with the system as it was only programmed to respond to pre-defined keywords from the participants. For instance, when they reported that they had reactions, the system would respond with an answer to instruct them on how to report without providing them any additional advice based on their reaction. They preferred to be given advice when they answered yes. **A female in her 20’s said:**

> “*For example, when I tell you that I have reactions, you are supposed to tell me that the medication that you are using is for a while but you will be okay. So, even as I take it I know that there is a day that I will be okay. However, when I say yes and then you keep quiet, on my side, the SMS is not good*.”

## Discussion

This paper investigated the feasibility and acceptability of using SMS messages to improve awareness about the importance of ADR reporting using both qualitative and quantitative approaches. Hence, we designed and developed an SMS system to trigger the active reporting of ADRs in Tanzania. Further, we explored the most common ADR reported by the participants.

Overall, we found that the majority of the participants who received the SMS question about whether they experienced ADR preferred to respond to the question. Furthermore, when asked if they wanted to report the symptoms, the majority were found to be willing to report the symptoms through the web-based route. Even though the majority responded to the question to favor the web-based tool, contrary to our expectations, the most used reporting strategy was clinic visits. Regarding experience, the majority expressed being satisfied with the SMS content and it was related to an increase in their willingness to report ADRs. Also, a large percentage of participants expressed being motivated by the SMS to report the ADR as the content of SMS was found to be well understood and easy to respond to. The two ways of communication were found to improve the interaction between the healthcare workers and participants. Furthermore, participants indicated a strong preference for the bi-weekly reminders as they felt to be cared for by someone.

A study from Kenya had a similar finding that a lower rate of ADRs was reported through the online route compared to reporting during the clinic visit(16). Another study by Mir et al reported that the reasons for participants not reporting online are based on the perception that no action will be taken if the report is sent web-based(17). A study in Ethiopia reported that personal contact with the study participants was associated with a higher number of reporting(18). A study in Ghana revealed that the disparity in financial status and poor internet connectivity throughout sub-Saharan Africa may lead to low rates of web-based reporting(19). Studies conducted in Europe and Asia have shown that the preferred means of reporting are email and website due to the low cost of the internet and stable internet infrastructure(19).

It was also noted in our study that the patients’ at the KCMC site, which is a referral hospital, more often reported ADR compared with Majengo health center. This could be due to a lack of awareness at the low facility levels. A study conducted in Ethiopia revealed insufficient knowledge of the existing PV system at health centers contributing to poor ADR reporting(7). Another study in Kenya described that the availability of pharmacists in big hospitals likely encourages reporting while health centers depend on nurses who have less knowledge about the PV systems (16). Hanafi S et al. suggests that a permanent ADR educational program is required for nurses in low-facility health centers to improve ADR reporting(20).

Moreover, in this study, we found that those who are on medication for more than 12 months report more compared to those on treatment for about three months. This suggests that being on treatment for a longer time might result in more ADR or more reporting and creates more awareness of ADRs. This is related to the previous study conducted in Ghana that showed that ART-experienced participants reported ADRs more frequently compared with those who just initiated the treatment (19).

In our study, we also saw that there are different ADRs reported from different patients. The same type of ADR profiles has been seen in other studies conducted in Tanzania, which concede our results(21). The reactions encountered from the drugs may be different for each participant due to different genetic differences from one person to another(22,23).

Our study has some limitations. First, this was a pilot study that was conducted within a short period in two health facilities and only a small sample size was included we also excluded participants who could not read. Therefore, the results from this study cannot be generalized to the entire community. Future studies should consider a larger sample size to investigate effectiveness. In addition, we do not know the acceptability and feasibility of the intervention if it is implemented for a longer duration of time.

Second, in this study, we could not confirm if the reported symptoms from the patients were actual side effects of the drugs or were due to disease progression. Tests like liver function, kidney function, and other cholesterol tests were not performed in this study. Therefore, future studies need to perform tests for patients who report symptoms.

It should be considered that this study has several strengths. In addition, this was a mixed-methods study, which used both quantitative and qualitative data in concluding the feasibility of using SMS service to report ADR in society.

Also, this study included both ART-naïve and treatment-experienced participants to tell if there could be differences in experiencing and reporting ADR. We collected data from both the participants and healthcare providers on their experiences of ADR reporting.

## Conclusion

In this study, we saw that using the common way of communication, which is SMS, could be a viable way to increase knowledge and understanding of the importance of reporting ADRs. In addition, it can improve communication between all people involved in the disease management of PLHIV to address the potential risks of ADRs. The use of SMS is a low-cost intervention and a simple way to improve public health issues within society. The participants in this study managed to use the SMS platform to report if they encountered any medication side effects. As this was a pilot feasibility study, the results may serve for larger studies to investigate the effect of the SMS system on ADR reporting and it may be scaled up to other regions and countries.

## Data Availability

Data for this manuscripts are fully available and can be shared upon request.

## Authors’ contributions

All authors have contributed equally during the development of the proposal, data collection procedures, analysis of the data, and writing and review of the manuscript.

## Acknowledgments

We appreciate all our study participants who consented to participate in our study. We also extend our sincere gratitude to all nurses and doctors at CTC sites, which were included in our study. We are grateful to the REMIND team at KCRI Tanzania for their endless efforts during the preparation and implementation of the study. Lastly, we thank the funder, the European and Developing Countries Clinical Trials Partnership for their support through the EACCR.

